# Intersecting vulnerabilities: Race, Depression, and White Matter Hyperintensity burden in Aging

**DOI:** 10.1101/2025.07.24.25332172

**Authors:** Farooq Kamal, Roqaie Moqadam, Cassandra Morrison, Mahsa Dadar

## Abstract

**BACKGROUND:** White matter hyperintensities (WMHs) are markers of brain aging and are associated with cognitive decline and dementia. However, research regarding how race, ethnicity, and depression status influence WMHs remains mixed. This study examined the interactive effects of race/ethnicity and depression on WMHs and cognition in older adults.

**METHODS:** Data from the National Alzheimer’s Coordinating Center included 2,411 older adults (773 Whites with Depression, 1,360 Whites without Depression, 89 Blacks with depression, 189 Blacks without depression). Bootstrap sampling (1,000 iterations) was used to match the White and Black samples. Linear regressions were then used to i) assess WMH differences across race/ethnicity and depression groups, and ii) to examine whether the associations between WMH burden and cognition were different across these groups.

**RESULTS:** Black older adults with depression showed greater global as well as regional WMH burden than Black older adults without depression (median *t* = 0.68–1.67), and depression significantly influenced the relationship between WMH burden and cognitive impairment in this group (median *t* = 1.15–2.03). Similar results were observed for Hispanics with depression (median *t* = 1.47–2.87), while WMH burden did not differ in White older adults with and without depression.

**CONCLUSIONS:** These findings suggest that race and depression may jointly influence cerebrovascular disease burden as well as its associations with cognition in aging and dementia.

## 1. Introduction

White matter hyperintensities (WMHs) are common markers of brain aging, reflecting damage to the white matter due to cerebral small vessel disease. They appear as bright spots on T2-weighted or fluid-attenuated inversion recovery (FLAIR) MRI scans or as hypointense regions in T1-weighted MRIs(1). WMHs are frequently observed in older adults, even those without clinically significant cognitive impairment(2). However, higher WMH burden is linked to an increased risk of cognitive decline, mild cognitive impairment (MCI), and dementia (e.g., Alzheimer’s disease, AD)(3–6). Typical indicators of increased WMH burden include vascular risk factors such as hypertension, obesity, and diabetes(7). However, other factors related to well-being such as depression have also been implicated in relation to higher WMH burden(8,9).

Depression frequently affects older adults, with 20% of the older adult population experiencing depression, a number that rises to above 40% for those in long-term care homes(10). Older adults with depression exhibit greater WMH burden compared to those without depression, which may play a role in their increased cognitive decline and development of dementia(11–13). More specifically, WMHs have been associated with depressive symptoms, with increased WMH burden in brain regions involved in mood and cognition, including the frontal, parietal, and temporal lobes(12–16). For example, the temporal lobes show increased WMH burden in late-life depression, with some studies linking these changes to emotional dysregulation and memory loss(13–16). To explain these findings, the vascular depression hypothesis suggests that cerebrovascular disease, as indexed by WMHs, may predispose or precipitate depression in older adults(15–16).

Prevalence of both WMHs and depression differs by race and ethnicity. Although some studies report no differences in WMH burden between racial groups(17), many show higher WMHs among Black compared to White individuals(18–20). More recently, several studies have observed that Black older adults exhibit greater WMH burden than White adults, but these differences are attenuated after adjusting for vascular risk factors(18,21,22). Furthermore, ethnicity differences in WMHs have also been observed, with Hispanic older adults showing distinct patterns of WMH distribution compared to non-Hispanic Whites(18,20,21,23).

Racial and ethnic differences have also been observed in depression. Similar to WMH findings, these results are also mixed. For example, a recent review observed that 31% of studies report that White older adults have more depression than Black older adults, 22% report that Blacks exhibit more depression than Whites, 22% show no race differences in depression, and 11% show mixed results(24). Beyond race, significant differences in depression severity have also been found between Hispanic and non-Hispanic older adults(25). In a large U.S. cohort, Hispanic participants exhibited a 23% higher overall depression severity compared to non-Hispanic Whites, even after controlling for sociodemographic, lifestyle, and health variables(25).

Although prior studies have examined the independent relationships between depression and WMHs as well as race/ethnicity and WMHs, less is known about whether the relationship between WMHs and depression varies by race. Given the limited research into the complex relationship between depression, WMHs, and cognition, examining these factors together may provide critical insights into racial disparities in aging. To address these gaps, the present study uses the National Alzheimer’s Coordinating Center (NACC) dataset to examine how depression, race (Black vs. White) and ethnicity (Hispanic vs. non-Hispanic) influence WMH burden in aging and AD. Our goal is to determine if depressed older adults show greater regional WMH burden compared to non-depressed older adults and whether these differences vary by race and ethnicity. Regional measures, especially in the frontal and parietal lobes, will be analyzed given their strong links to small vessel disease and AD, respectively(26–28).

## 2. Methods and Materials

### 2.1 National Alzheimer’s Coordinating Center

The data for this study was obtained from the National Alzheimer’s Coordinating Center (NACC, https://naccdata.org/) database, and included the NACC Uniform Data Set (UDS), and MRI Data Set(29–31). Participants were selected from the NACC database based on availability of clinical data and MRI scans for extracting WMH measurements. With regards to race, there were a total of 2411 participants comprising 1,360 White participants with no depression, 773 White participants with depression, 189 Black participants with no depression, and 89 Black participants with depression.

When examining ethnicity, the dataset comprised a total of 2302 participants consisting of 1368 non-Hispanics with no depression, 116 Hispanics with no depression, 731 non-Hispanics with depression, and 87 Hispanics with depression.

Clinical diagnoses for each participant were determined using two variables from the NACC dataset: NACCETPR and NACCTMCI. The NACCETPR variable indicates the primary etiologic diagnosis assigned by clinicians, identifying individuals with AD etiology or no cognitive impairment. Among participants who did not meet criteria for AD and were not classified as cognitively normal, the NACCTMCI variable was used to identify MCI based on clinical evaluation. Participants with other diagnoses (e.g. Parkinson’s disease, Lewy body dementia, frontotemporal dementia) were excluded.

Depression status was determined using multiple indicators from the NACC dataset. Depression status was coded as a binary variable (1=depressed, 0=not depressed). Participants were coded as depressed if they had a presumptive etiologic diagnosis of depression (DEP), reported depression or dysphoria in the past month (DEPD), or had active depression in the last two years (DEP2YRS). Additionally, individuals with a primary etiologic diagnosis of depression (NACCETPR=19) were also coded as depressed. Participants were coded as not depressed if they had no indication of depression across these variables.

### 2.2 Vascular Risk Factors

Body Mass Index (BMI) was derived from the NACCBMI variable, which is based on participants’ recorded height and weight during their visit. Hypertension status was determined by combining information from the HYPERT, HXHYPER, and HYPERTEN variables in the NACC dataset, with individuals coded as ‘1’ if hypertension was present and ‘0’ if absent. Similarly, diabetes status was assessed using the DIABET and DIABETES variables, where a value of ‘1’ indicated a diagnosis of diabetes and ‘0’ indicated no diabetes.

### 2.3 Volumetric WMH Measurements

T1-weighted (T1w) MRI scans were pre-processed using our established pipeline, which included noise reduction(32), intensity inhomogeneity correction(33), and intensity normalization into range [0-100]. The pre-processed resulting images were then linearly (9 parameters: 3 translation, 3 rotation, and 3 scaling) registered to the MNI-ICBM152-2009c average(34,35).

WMH measurements were extracted using a previously validated automated segmentation method(35,36). This technique has been used in other multi-center studies(12,37) as well as in NACC cohorts(12,38). The automated WMH segmentation technique extracts a set of location (i.e. spatial priors) and intensity (distribution histograms) features and uses them in combination with a random forest classifier to detect the WMHs in new images(33,34,39). Automatic segmentation of the WMHs was completed using only the T1w contrasts as FLAIR scans were not consistently available across participants in the NACC dataset. The accuracy of this T1w-based approach has been validated in earlier work, demonstrating a strong correlation with WMH volumes derived from FLAIR scans (*r*=.97,*p*<.001)(33,34,39). All preprocessing steps and WMH segmentations were visually reviewed by an experienced rater (RM) blinded to clinical diagnosis. WMH burden was quantified as the total volume of voxels identified as WMH in standard space (measured in mm³) and normalized for head size. Total and regional (frontal, temporal, occipital, and parietal) WMH volumes were calculated based on Hammers Atlas(35,40). All WMH volumes were log-transformed to obtain a normal distribution.

### 2.4 Statistical Analysis

All analyses were conducted in MATLAB R2021a. Demographic group differences were assessed using independent sample t-tests for continuous variables including age, education, and BMI, and chi-square (*x*^2^) tests for categorical measures including sex, hypertension, and diabetes. Older adults were categorized into four groups for race-by-depression comparisons: White with depression, White without depression, Black with depression, and Black without depression. For ethnicity-by-depression comparisons, participants were grouped as Hispanic with depression, Hispanic without depression, non-Hispanic with depression, and non-Hispanic without depression. Race-by-depression and ethnicity-by-depression groups were treated as categorical predictors. Linear regression models were utilized to explore the differences in total and regional WMH burden (frontal, parietal, temporal, and occipital lobes) across depression and race/ethnicity groups. More specifically, WMH burden was modeled as a function of race and depression and adjusted for age, sex, education, and baseline diagnostic status. Diagnosis was included as a categorical covariate in the model to account for potential diagnostic differences across racial groups.

*WMH ∼ Race-Depression/Ethnicity-Depression+Age+Sex+Education+Diagnosis (1)*

To evaluate the role of vascular risk factors, additional models were assessed that included diabetes, hypertension, and BMI as covariates. These analyses were conducted to examine whether any race or ethnicity and depression group differences were driven by differences in vascular risk factors known to influence WMH burden.

*WMH ∼ Race-Depression/Ethnicity-Depression+Age+Sex+Education+Diagnosis+Diabetes+Hypertension+BMI (2)*

To examine whether the association between WMH and cognition differed across race and depression groups, a linear regression model was completed, with cognition as dependent variable and total or regional WMH burden as predictors. Cognition was measured using Clinical Dementia Rating Sum of Boxes (CDR-SB). WMH burden was examined across five regions: total, frontal, temporal, parietal, and occipital. Covariates included age, years of education, and baseline diagnostic status. The model included an interaction term between Race × Depression (White no depression, White with depression, Black no depression, Black with depression) and WMH, adjusting for age, sex, education, and diagnostic status.

*Cognition ∼ Race-Depression/Ethnicity-Depression × WMH+Age+Sex+Education+Diagnosis (3)*

To account for group imbalances in number and age, sex, education, and diagnostic status between White and Black/Hispanic participants, a bootstrapping approach was implemented. This method involved repeatedly sampling the larger comparison groups (e.g., White and non-Hispanic participants) to create subsamples that were matched to the smaller groups (e.g., Black and Hispanic participants) on age, sex, education, and diagnosis(40). In each iteration, a subset of White participants was randomly selected to match the size and characteristics of the smaller group being compared. For example, in comparisons between racial groups, White individuals were randomly drawn from the original sample to mirror the distribution of the Black group across age, sex, education, diagnosis. This resampling procedure was repeated 1,000 times to generate balanced samples for each racial and ethnic comparison. The same process was applied to comparisons involving Hispanics, ensuring equivalent sample sizes and covariate distributions. This strategy follows established best practices for addressing imbalanced data in large-scale population studies(22,40). The bootstrapping and resampling process was applied consistently for all models, and median *t-*statistic, 95% confidence intervals (CI, corresponding to a *p*-value of .05), and 99.5% CI (corresponding to a *p*-value of .005) were reported. All continuous variables were standardized (z-scored) across the sample prior to conducting regression and mediation analyses. Linear regression models were implemented using the fitlm function.

## 3. Results

### 3.1 Demographic and Vascular risk factors

Table 1 presents the demographic and vascular risk factor information across racial and depression groups. White participants with depression were significantly younger (*t=*-2.88, *p*=.004), had lower education levels (*t=*-3.65, *p*<.001), and were more likely to have diabetes (*x²*=5.59, *p*=.018) than White participants without depression. There were no significant group differences in sex, BMI, or hypertension between these groups. Among Black participants, there were no significant differences between those with and without depression across age, sex, education, BMI, hypertension, or diabetes. When comparing White and Black participants with depression, White participants had significantly higher education levels (*t=*2.78, *p*=.006), more males (x²=3.88, p=.049), lower BMI (*t=*-4.40, *p*<.001), and lower rates of hypertension (*x²*=34.05, *p*<.001) and diabetes (*x²=*25.42, *p*<.001) than Black participants with depression. When comparing White and Black participants without depression, White participants had higher education levels (*t=*4.74, *p*<.001), more males (*x*²=21.54, *p*<.001), lower BMI (*t=*-6.81, *p*<.001), and lower rates of hypertension (*x*²=48.23, *p*<.001) and diabetes (*x*²=99.26, *p*<.001) than Black participants.

**Table 1.**
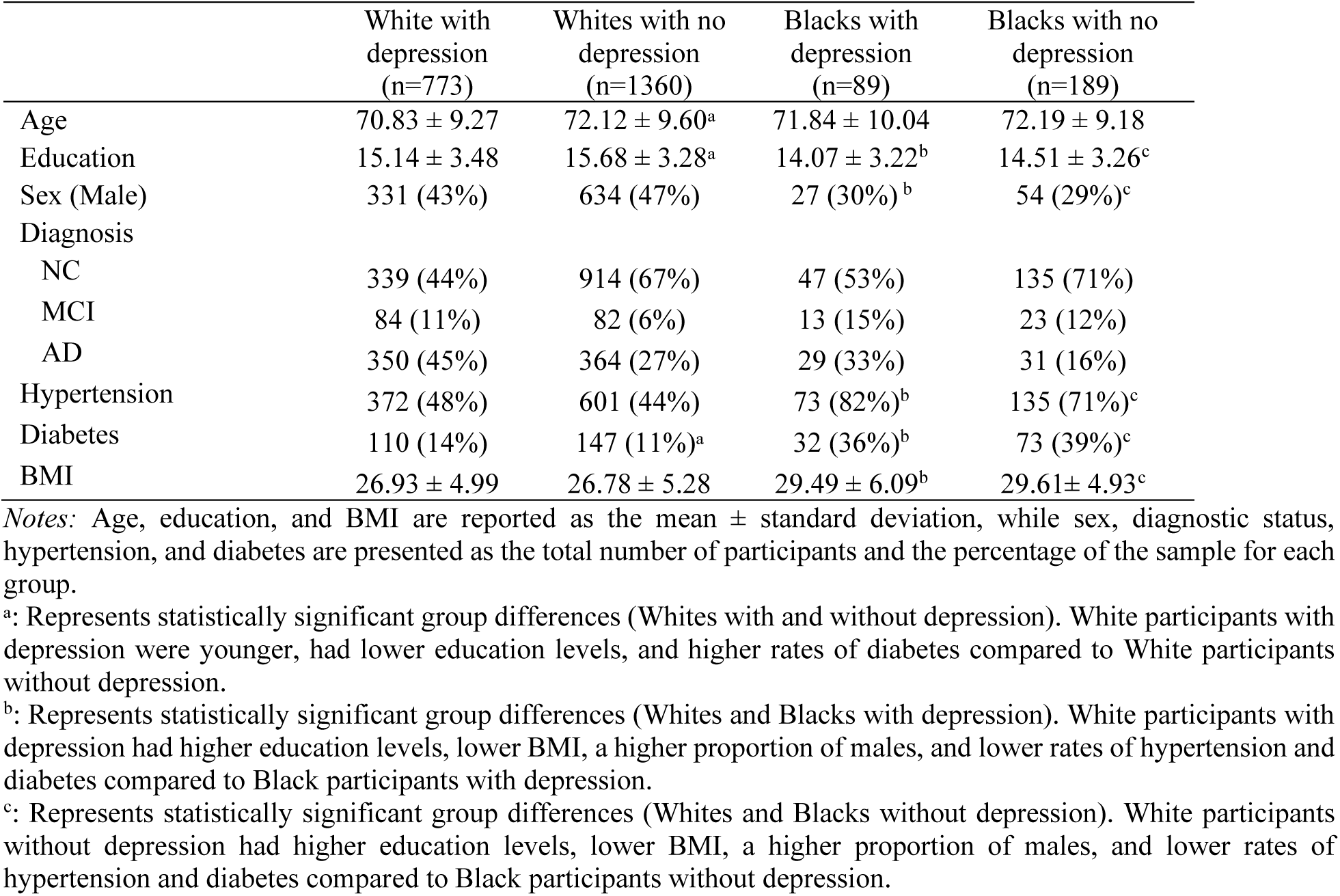
Demographic information for vascular risk factors by race and depression.

Table 2 presents the demographic and vascular risk factor information across ethnic and depression groups. Hispanic participants with and without depression did not significantly differ in age, sex, education, BMI, hypertension, or diabetes. In contrast, non-Hispanic participants with depression were significantly older (*t=*2.87, *p*=.004) and had lower education levels (*t=*-3.48, *p*<.001), and lower rates of hypertension (*x*²=4.25, *p*=.039) than non-Hispanics without depression, though no significant differences were observed in sex, BMI, or diabetes. Hispanic participants with depression were older (*t*=2.13, *p*=.034), lower education levels (*t*=-11.34, *p*<.001), fewer males (*x*²=-7.98, *p*=.004), and lower rates of hypertension (*x*²=-5.50, *p*=.02), and diabetes (*x*²= - 21.91, *p*<.001) than non-Hispanic participants with depression. Similarly, Hispanic participants without depression were older (*t=*2.26, *p*=.024) and had significantly lower education (*t=*-15.82, *p*<.001), had more males (*x*²=5.29, *p*=.021), and exhibited higher rates of hypertension (*x*²=18.57, *p*<.001), and diabetes (*x*²=24.14, *p*<.001), compared to non-Hispanics without depression.

**Table 2.**
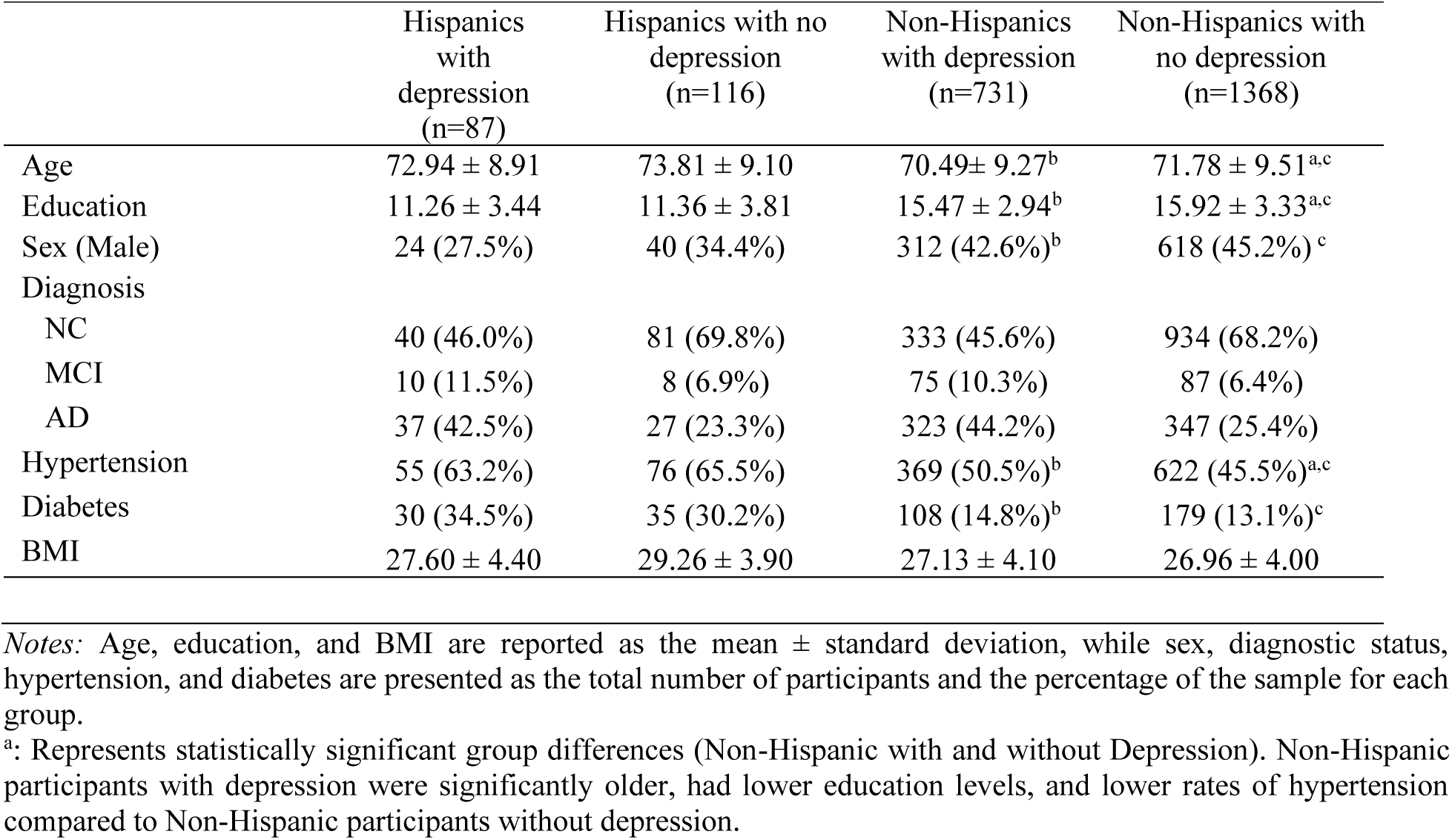
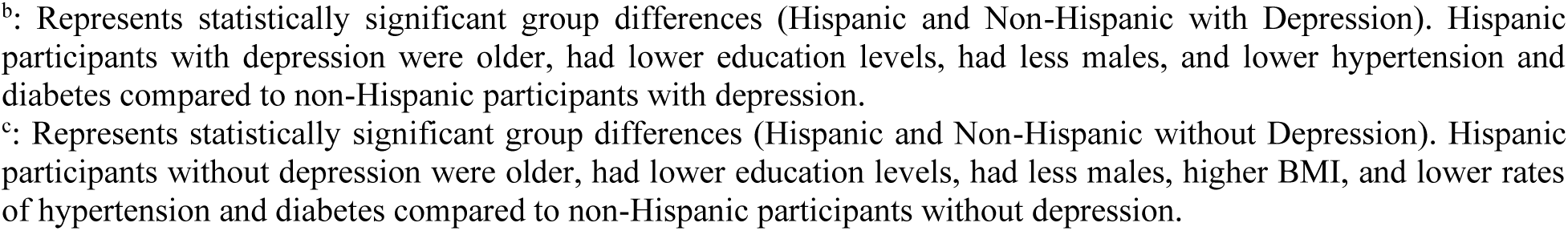
Demographic information for vascular risk factors by ethnicity and depression.

|  | Hispanics<br>with<br>depression<br>(n=87) | Hispanics with no<br>depression<br>(n=116) | Non-Hispanics<br>with depression<br>(n=731) | Non-Hispanics with<br>no depression<br>(n=1368) |
| --- | --- | --- | --- | --- |
| Age | 72.94 ± 8.91 | 73.81 ± 9.10 | 70.49 ± 9.27<sup>b</sup> | 71.78 ± 9.51<sup>a,c</sup> |
| Education | 11.26 ± 3.44 | 11.36 ± 3.81 | 15.47 ± 2.94<sup>b</sup> | 15.92 ± 3.33<sup>a,c</sup> |
| Sex (Male) | 24 (27.5%) | 40 (34.4%) | 312 (42.6%)<sup>b</sup> | 618 (45.2%)<sup>c</sup> |
| Diagnosis |  |  |  |  |
| NC | 40 (46.0%) | 81 (69.8%) | 333 (45.6%) | 934 (68.2%) |
| MCI | 10 (11.5%) | 8 (6.9%) | 75 (10.3%) | 87 (6.4%) |
| AD | 37 (42.5%) | 27 (23.3%) | 323 (44.2%) | 347 (25.4%) |
| Hypertension | 55 (63.2%) | 76 (65.5%) | 369 (50.5%)<sup>b</sup> | 622 (45.5%)<sup>a,c</sup> |
| Diabetes | 30 (34.5%) | 35 (30.2%) | 108 (14.8%)<sup>b</sup> | 179 (13.1%)<sup>c</sup> |
| BMI | 27.60 ± 4.40 | 29.26 ± 3.90 | 27.13 ± 4.10 | 26.96 ± 4.00 |

Notes: Age, education, and BMI are reported as the mean ± standard deviation, while sex, diagnostic status, hypertension, and diabetes are presented as the total number of participants and the percentage of the sample for each group.
<sup>a</sup>: Represents statistically significant group differences (Non-Hispanic with and without Depression). Non-Hispanic participants with depression were significantly older, had lower education levels, and lower rates of hypertension compared to Non-Hispanic participants without depression.
<sup>b</sup>: Represents statistically significant group differences (Hispanic and Non-Hispanic with Depression). Hispanic participants with depression were older, had lower education levels, had less males, and lower hypertension and diabetes compared to non-Hispanic participants with depression.
<sup>c</sup>: Represents statistically significant group differences (Hispanic and Non-Hispanic without Depression). Hispanic participants without depression were older, had lower education levels, had less males, higher BMI, and lower rates of hypertension and diabetes compared to non-Hispanic participants without depression.

### 3.3 WMH differences due to depression and race

Figures 1 and 2 show violin plots of total and regional WMH volumes by race and depression, and ethnicity and depression groups, respectively. Figure 3 provides an example of regional WMH segmentations for a NACC participant. Table 3 presents the bootstrapped median t-statistics and 95% confidence intervals comparing race and depression groups on total and regional WMH burden before controlling for risk factors. Linear regression analyses (Model 1) revealed significant race and some depression effects for both total and regional WMH volumes. Black older adults without depression showed significantly higher total (*t*=4.64), frontal (*t*=4.08), parietal (*t*=5.40), temporal (*t*=2.33), and occipital (*t*=4.44) WMH burden than White older adults without depression. Black older adults with depression had higher total (*t*=3.46), frontal (*t*=3.42), parietal (*t*=3.52), and occipital (*t*=2.79) WMH burden than White older adults with depression, while temporal WMH did not differ between the groups (*t*=1.08). Total or regional WMH burden did not differ in Whites with and without depression. Blacks with depression exhibited higher total (*t*=1.25), frontal (*t*=1.67), and parietal (*t*=0.68) WMH burden compared to Blacks without depression, with temporal (*t*= –0.10) and occipital (*t*= –0.17) regions showing no significant differences linked to depression.

**Figure 1.**
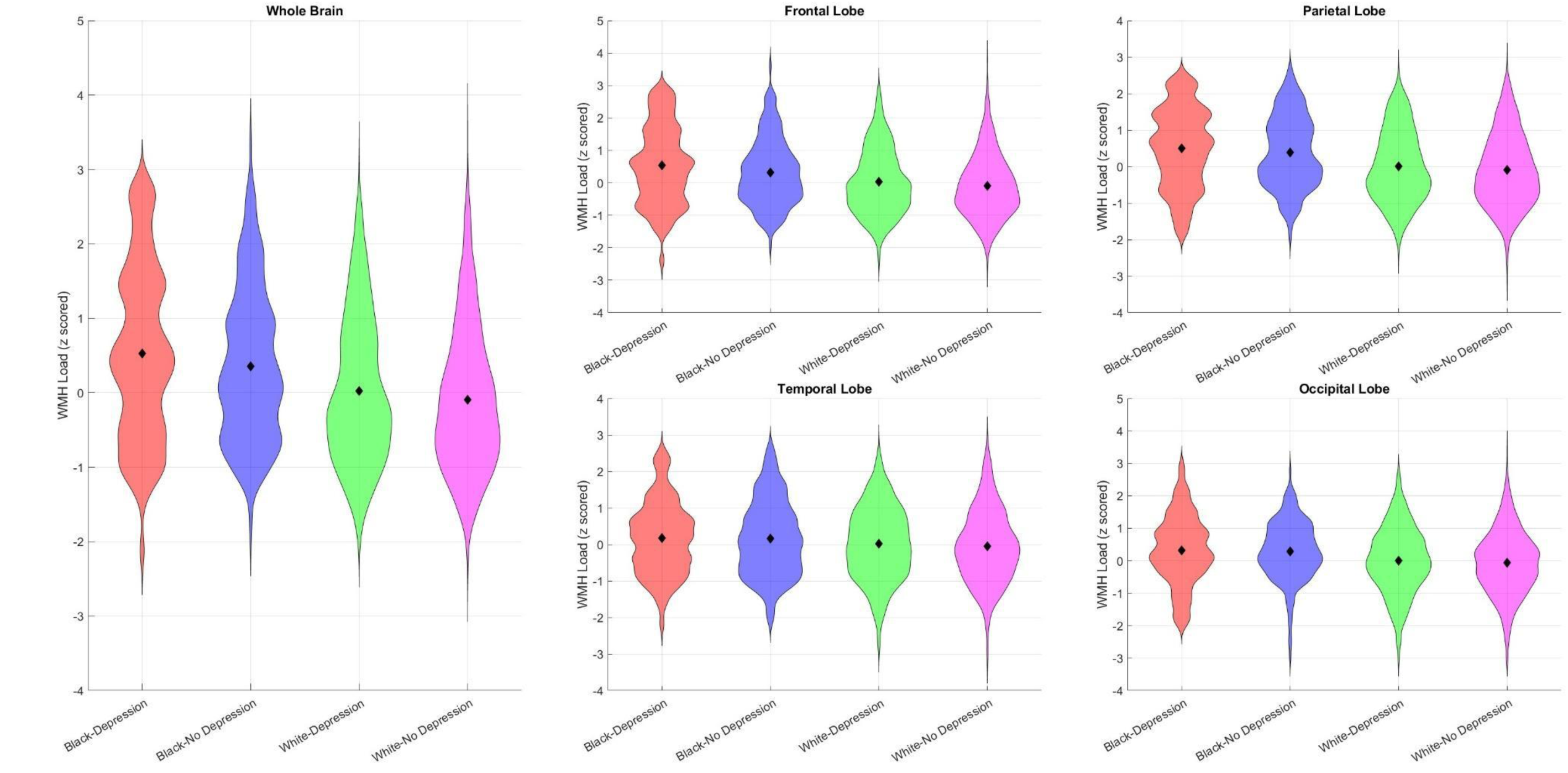
Violin plots show total and regional WMH distributions (log transformed and normalized) across race and depression groups. *Notes:* The shape of each violin reflects the smooth distribution of variability and spread. Black diamond markers denote group means. The width of the plots indicates the relative density of participants at different values. WMH = white matter hyperintensity.

**Figure 2.**
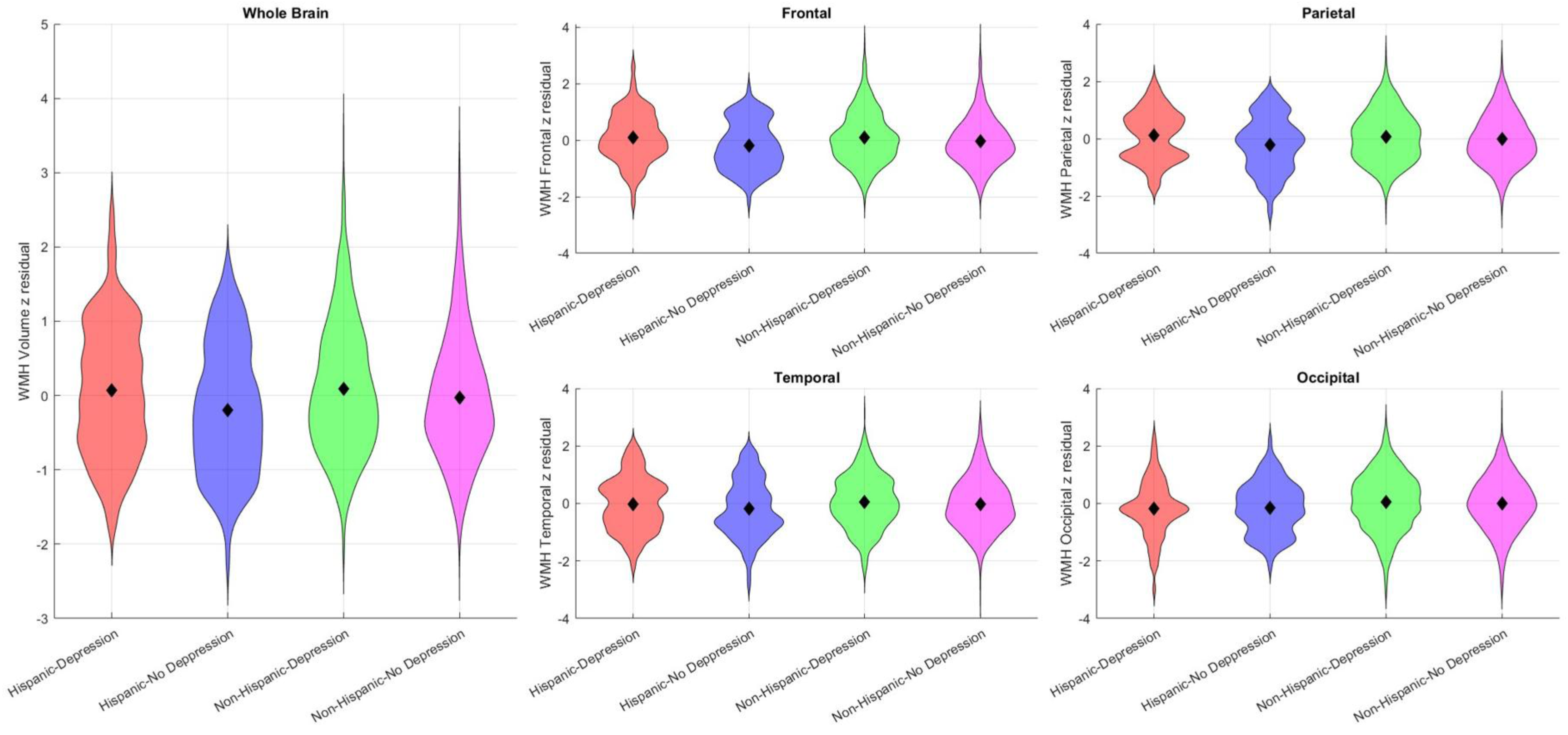
Violin plots show baseline total and regional WMH distributions (log transformed and normalized) across ethnic and depression groups. *Notes:* The shape of each violin reflects the smooth distribution of variability and spread. Black diamond markers denote group means. The width of the plots indicates the relative density of participants at different values. WMH = white matter hyperintensity.

**Figure 3.**
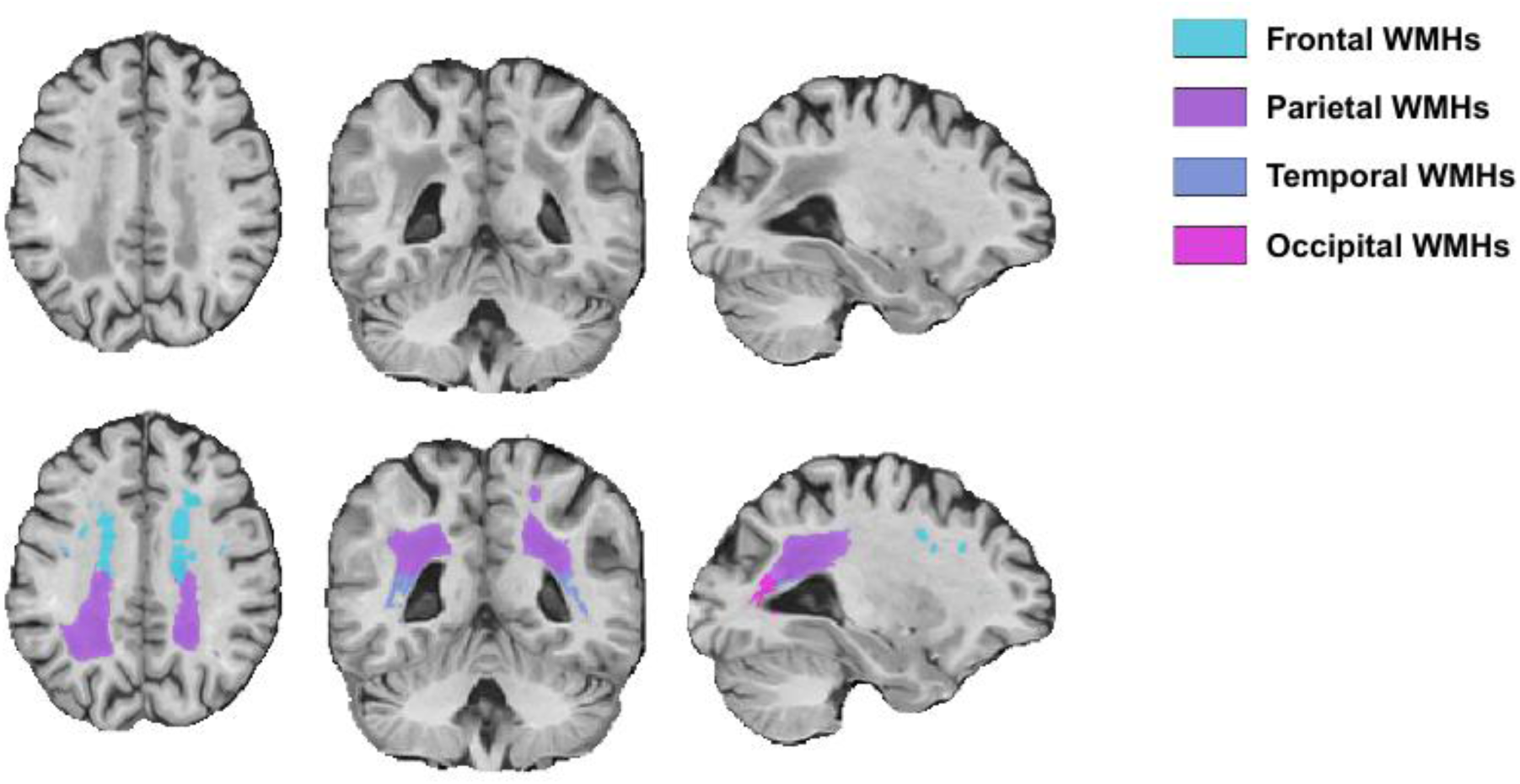
Example of volumetric WMH segmentation of a NACC participant.

**Table 3:** Median t-statistic (95% CI) across 1,000 iterations comparing race and depression across total and regional WMH burden before controlling for risk factors (Model 1).

| *Comparison* | *Total WMH* | *Frontal WMH* | *Parietal WMH* | *Temporal WMH* | *Occipital WMH* |
| --- | --- | --- | --- | --- | --- |
|  | *Median T stat*<br>*[95% CI]* | *Median T stat*<br>*[95% CI]* | *Median T stat*<br>*[95% CI]* | *Median T stat*<br>*[95% CI]* | *Median T stat*<br>*[95% CI]* |
| White with Depression vs<br>White with no Depression | 1.17<br>[-0.64, 2.97] | 1.20<br>[-0.48, 2.89] | 1.14<br>[-0.70, 3.02] | 0.50<br>[-1.26, 2.53] | 0.30<br>[-1.57, 2.18] |
| Black with no Depression vs<br>White with no Depression | **4.64**<br>**[3.39, 5.93]\*** | **4.08**<br>**[2.79, 5.44]\*** | **5.40**<br>**[3.93, 6.84]\*** | **2.33**<br>**[1.08, 3.73]\*** | **4.44**<br>**[3.09, 5.82]\*** |
| Black with Depression vs<br>White with Depression | **3.46**<br>**[2.17, 4.67]\*** | **3.42**<br>**[2.11, 4.77]\*** | **3.52**<br>**[2.15, 4.89]\*** | 1.08<br>[-0.29, 2.44] | **2.79**<br>**[1.40, 4.13]\*** |
| Black with Depression vs<br>Black with no Depression | **1.25**<br>**[1.03, 1.45]\*** | **1.67**<br>**[1.46, 1.87]\*** | **0.68**<br>**[0.49, 0.89]\*** | -0.10<br>[-0.30, 0.10] | -0.17<br>[-0.40, 0.04] |

*Note. An asterisk (\*) marks contrasts whose bootstrapped 99.5 % confidence interval excludes zero, indicating significance even after the stricter threshold.*

Table 4 shows the median t-statistics and 95% confidence intervals for Ethnicity and depression contrasts across total and regional WMH burden before controlling for risk factors. Linear regression analyses (Model 1) revealed significant ethnicity and depression effects for both total and regional WMH volumes. Hispanics without depression showed significantly higher total (*t*=2.11), frontal (*t*=2.02), parietal (*t*=2.27) and temporal (*t=*1.78), WMH burden compared to Non-Hispanics without depression. Non-Hispanics with depression exhibited significantly greater temporal (*t*=1.49) and occipital WMH burden (*t*=2.32) than Hispanics with depression, although total, frontal and parietal WMHs did not significantly differ. However, only occipital WMH burden was significant at 99.5% CI. Hispanics with depression had greater total (*t*=2.69), frontal (*t*=2.86), parietal (*t*=2.87), and temporal (*t*=1.47) WMH than their Hispanics with no depression in both models, with no difference in the occipital regions. Non-Hispanics with and without depression did not differ in total or regional WMH burden.

**Table 4:** Median t-statistic (95% CI) across 1,000 iterations comparing race and depression across total and regional WMH burden before controlling for risk factors (Model 1).

| *Comparison* | *Total WMH* | *Frontal WMH* | *Parietal WMH* | *Temporal WMH* | *Occipital WMH* |
| --- | --- | --- | --- | --- | --- |
|  | *Median T stat*<br>*[95% CI]* | *Median T stat*<br>*[95% CI]* | *Median T stat*<br>*[95% CI]* | *Median T stat*<br>*[95% CI]* | *Median T stat*<br>*[95% CI]* |
| Non-Hispanic with no<br>Depression vs Hispanic with<br>no Depression | **2.11\***<br>**[0.70, 3.42]** | **2.02**<br>**[0.67, 3.37]** | **2.27\***<br>**[0.86, 3.60]** | **1.78**<br>**[0.56, 3.16]** | 1.18<br>[-0.23, 2.60] |
| Non-Hispanic with<br>Depression vs Hispanic with<br>Depression | 0.52<br>[- 0.74, 1.81] | 0.23<br>[-1.12, 1.55] | 0.56<br>[-0.70, 1.97] | **1.49**<br>**[0.32, 2.69]** | **2.32\***<br>**[ 0.99, 3.62]** |
| Hispanic with Depression vs<br>Hispanic with no Depression | **2.69\***<br>**[2.40, 3.01]** | **2.86\***<br>**[2.54, 3.18]** | **2.87\***<br>**[2.57, 3.17]** | **1.47\***<br>**[1.20, 1.73]** | -0.01<br>[-0.28, 0.25] |
| Non-Hispanic with<br>Depression vs Non-Hispanic<br>with no Depression | 1.29<br>[-0.52, 3.19] | 1.16<br>[-0.66, 3.04] | 1.23<br>[-0.45, 3.18] | 1.29<br>[-0.39, 3.10] | 1.30<br>[-0.66, 3.19] |

*Note. An asterisk (\*) marks contrasts whose bootstrapped 99.5 % confidence interval excludes zero, indicating significance even after the stricter threshold.*

In the models controlling for vascular risk factors (Model 2), the race by depression interaction effects remained the same in terms of significance and effect size. For the ethnicity by depression models, almost all results remained the same, however, the difference between Hispanics and non-Hispanics with depression in the temporal lobe were no longer significant.

### 3.4 Association between cognition and WMH

To examine whether the association between WMH and cognition (CDR-SB score) differed across race/ethnicity and depression status, a linear regression model (Model 3) was completed. Table 5 shows the median t-statistics and 95% confidence intervals for the interaction term between race and depression groups and WMHs, reflecting the group differences in the impact of WMH burden on cognition. The results indicated that Black older adults with depression had a significantly stronger positive association between WMH burden and CDR-SB scores than White older adults with depression. This effect was observed across total WMH (*t*=1.55), frontal (*t*=1.46), and parietal (*t*=1.42) regions. However, these associations did retain statistical significance with the more strict 99.5% threshold.

**Table 5.** Median t-statistic (95% CI) across 1,000 iterations comparing race and depression across total and regional WMH burden on cognition.

| *Comparison* | *Total WMH* | *Frontal WMH* | *Parietal WMH* | *Temporal WMH* | *Occipital WMH* |
| --- | --- | --- | --- | --- | --- |
|  | *Median T stat*<br>*[95% CI]* | *Median T stat*<br>*[95% CI]* | *Median T stat*<br>*[95% CI]* | *Median T stat*<br>*[95% CI]* | *Median T stat*<br>*[95% CI]* |
| White with Depression vs<br>White with no Depression | -0.73<br>[-2.60, 1.36] | -0.52<br>[-2.50, 1.59] | -0.70<br>[-2.66, 1.43] | -0.58<br>[-2.41, 1.44] | -0.75<br>[-3.12, 1.79] |
| Black with no Depression vs<br>White with no Depression | -0.92<br>[-2.49, 0.45] | -0.46<br>[-2.08, 1.05] | -1.05<br>[-2.47, 0.25] | **-1.39**<br>**[-2.83, -0.11]** | **-1.89**<br>**[-3.85, -0.38]** |
| Black with Depression vs<br>White with Depression | **1.55**<br>**[0.01, 3.02]** | **1.46**<br>**[0.03, 2.92]** | **1.42**<br>**[0.02, 2.97]** | 1.27<br>[-0.16, 2.66] | 0.20<br>[-1.34, 1.78] |
| Black with Depression vs<br>Black with no Depression | **1.85**<br>**[1.51, 2.17]\*** | **1.56**<br>**[1.27, 1.86]\*** | **1.86**<br>**[1.58, 2.20]\*** | **2.03**<br>**[1.72, 2.38]\*** | **1.15**<br>**[0.91, 1.40]\*** |

*Note.* An asterisk (\*) marks contrasts whose bootstrapped 99.5 % confidence interval excludes zero, indicating significance even after the strictest correction.

Additionally, White older adults without depression had a significantly stronger negative association between WMH burden and CDR-SB scores compared to White older adults without depression. This effect was observed in the temporal (*t*= –1.39) and occipital (*t*= –1.89) regions, but did not remain significant with the more strict 99.5% threshold. The analyses also indicated that Black older adults with depression had a significantly stronger positive association between WMH burden and cognitive scores than Black older adults without depression. This effect was observed across total WMH (*t*=1.85), frontal (*t*=1.56), parietal (*t*=1.86), temporal (*t*=2.03), and occipital (*t*=1.15) regions, all of which also remained significant with the 99.5% threshold. These findings suggest that among Black older adults, higher WMH burden is more strongly linked to greater cognitive dysfunction in those with depression than those without depression. For White older adults, the relationship between WMH and cognition did not significantly differ between those with and without depression.

Table 6 summarizes the results for the ethnicity/depression groups. Relative to Hispanics with depression, Non-Hispanics with depression showed a significantly weaker association between WMHs and cognition for total (*t*= –1.51) and frontal lobe (*t*= –1.80) WMHs. Only the frontal lobe association remained significant with the 99.5% threshold. No other group comparisons yielded significant interactions.

**Table 6.** Median t-statistic (95% CI) across 1,000 iterations comparing ethnicity and depression across total and regional WMH burden on cognition.

| *Comparison* | *Total WMH* | *Frontal WMH* | *Parietal WMH* | *Temporal WMH* | *Occipital WMH* |
| --- | --- | --- | --- | --- | --- |
|  | *Median T stat*<br>*[95% CI]* | *Median T stat*<br>*[95% CI]* | *Median T stat*<br>*[95% CI]* | *Median T stat*<br>*[95% CI]* | *Median T stat*<br>*[95% CI]* |
| Non-Hispanic with no<br>Depression vs Hispanic with<br>no Depression | 0.15<br>[-0.79, 1.33] | 0.71<br>[-0.22, 1.81] | -0.03<br>[-1.01, 1.07] | -0.43<br>[-1.39, 0.72] | -1.30<br>[-2.38, 0.02] |
| Non-Hispanic with<br>Depression vs Hispanic with<br>Depression | **-1.51**<br>**[-2.52, -0.35]** | **-1.80\***<br>**[-2.85, -0.86]** | -0.79<br>[-1.92, 0.15] | -1.15<br>[-2.26, 0.01] | 0.27<br>[-0.91, 1.63] |
| Hispanic with Depression vs<br>Hispanic with no Depression | 0.25<br>[-0.14, 0.37] | 1.03<br>[-0.88, 1.16] | -0.11<br>[-0.19, 0.01] | -0.29<br>[-0.43, 0.16] | -1.32<br>[-2.14, 1.79] |
| Non-Hispanic with<br>Depression vs Non-Hispanic<br>with no Depression | -0.25<br>[-2.03, 1.11] | -0.60<br>[-3.24, 0.19] | -0.98<br>[-2.45, 0.61] | -1.13<br>[-2.68, 0.34] | -0.49<br>[-2.19, 1.12] |

*Note.* An asterisk (\*) marks contrasts whose bootstrapped 99.5 % confidence interval excludes zero, indicating significance even after the strictest correction.

## 4. Discussion

The current study examined (i) whether WMH burden differs across race/ethnicity by depression groups and (ii) whether the association between WMH burden and cognition varies across those same groups. Our findings showed significant differences across race/ethnicity and depression groups for both WMH burden and its relationship with cognition. For Black older adults, depression was associated with higher total, frontal, and parietal WMH burden compared to Black older adults without depression. White older adults with depression showed no significant differences in WMH burden compared to those without depression. Within Hispanics, depression was associated with higher total, frontal, parietal, and temporal WMH, whereas occipital WMH burden remained unchanged. In contrast, WMH differences between depressed and non-depressed Non-Hispanics did not reach significance in any region. Hispanics and Non-Hispanics differed only in temporal and occipital WMH, with Non-Hispanics with depression showing higher WMH burden than Hispanics with depression. Additionally, the relationship between WMHs, depression, and cognition was stronger in Blacks and Hispanics than in Whites and Non-Hispanics. Importantly, the inclusion of vascular risk factors (diabetes, hypertension, BMI) did not significantly change the race by depression effects on WMH burden, suggesting that these group differences may not be solely driven by traditional vascular risk factors.

Regardless of depression status, Black older adults had more WMH burden compared to White older adults, supporting previous findings that Black older adults carry a disproportionately high cerebrovascular disease burden(14,18–22). Racial differences in WMH burden further differed based on depression status. Although depression was not associated with increased WMH burden in White older adults, Blacks with depression had increased burden compared to Blacks without depression. These findings align with prior work showing high WMH accumulation in Black adults with depressive symptoms(14). This increase, particularly in frontal WMHs in Blacks with depression is consistent with previous research highlighting the role of fronto-subcortical regions in late-life depression(41). The other regional differences (temporal, parietal and total WMH burden) are also central to mood regulation with WMH burden in these regions commonly linked to depressive symptoms in late life(12–16). Furthermore, Boots et al.(42) found that inflammatory markers associated with cerebrovascular risk were related to reduced WMH specifically in Black older adults, reinforcing the link between inflammation, WMH, and depression in this group. Our findings extend this literature by showing that WMHs is heightened in Black older adults with depression, indicating that cerebrovascular pathology may be a key mechanism linking depression and brain aging.

In addition to influencing vascular pathology, this study observed that depression interacted with WMHs to influence cognition. Previous research has shown that WMH burden is associated with cognitive impairment and dementia risk in older adults(3–5,43). The stronger link between depression and WMHs in Black compared to White older adults was further exemplified by the significant interaction between depression and WMHs impacting their cognition. Among older adults with depression, Black older adults showed a significantly steeper relationship between WMH (total, frontal, parietal) and poorer global cognition than their White counterparts. Furthermore, among Black older adults, depression significantly strengthened the association between WMH burden and cognitive decline across total, frontal, and parietal, temporal, and occipital regions, a finding that was not observed among White older adults. Together, these findings indicate that depression may amplify the cognitive effects of WMH burden in Black older adults. This racial difference is consistent with studies reporting higher cumulative vascular and neurodegenerative burden in minoritized groups when comorbid conditions such as depression are present(20,44).

Several factors might explain the greater impact of depression on WMH burden and their association with cognition in Black older adults. First, the White older adults had higher levels of education and may benefit from greater cognitive reserve compared to the Black older adults, allowing comparable WMH loads to exert smaller observable effects on cognition. Additionally, according to the vascular depression hypothesis, small-vessel disease may contribute to depressive symptoms only once a critical threshold is reached(41,45). Consistent with the vascular depression hypothesis, our results suggest that in Black older adults, depression may represent a cerebrovascular risk factor in which small-vessel disease contributes both to cognitive impairment and mood disorder. Conversely, the lack of significant depression effects in White older adults may reflect greater heterogeneity in the underlying role of depression, with vascular contributions playing a smaller role. It is possible that the neurobiological associations may differ across racial groups. In particular, chronic stress associated with lifelong experiences of racial discrimination might amplify depression-related cerebrovascular changes among Black older adults(20,46). Depression in older Black adults may also be influenced by factors in addition to cerebrovascular damage, such as socioeconomic stress or limited healthcare access(25,47). Overall, these group differences underscore the need to investigate non-vascular factors such as perceived discrimination, chronic stress, and socioeconomic barriers to better understand how depression and race together shape brain health.

Differences in the associations between depression, WMHs, and cognition were also observed when comparing Hispanic and Non-Hispanic older adults. When comparing depressed Hispanic and non-Hispanic older adults, depressed Non-Hispanics showed greater WMH than Hispanic older adults in the temporal and occipital regions. This finding is surprising given the higher late-life depression severity often reported in Hispanic compared to Non-Hispanic adults(25). According to the vascular depression hypothesis, this finding implies that small-vessel disease may underlie depression in ethnic groups, but baseline vascular health and non-vascular resilience factors such as social support likely moderate the impact on WMHs(14,45). Interestingly, the interaction between WMHs and depression on cognition was stronger in Hispanics compared to Non-Hispanics with depression, particularly in total and frontal WMHs. One possible interpretation (similar to the White adults) is that Non-Hispanics may be more resilient to WMH burden than Hispanic older adults. Additionally, in Hispanics, small increases in WMHs may indicate greater vascular vulnerability than in Non-Hispanics or they may have other modifiers that increase the cognitive impact (e.g., vascular risk factors or social determinants of health). Future research should explore the mediating mechanisms associated with these findings.

A key strength of this work is the use of a large, well-characterized dataset with robust measures of WMH burden. The bootstrapping approach employed is also a strength, supporting the validity of our results. Still, several limitations should be noted. First, NACC participants often differ in education and health status from the general population therefore may not be generalizable or representative(48). This study also relied on broad racial and ethnic categories, which can mask within-group differences(44). Third, the analyses presented did not measure depression severity, duration, early vs late onset of depression over time, factors that could provide more detail about how depression influences brain structure.

Overall, the absence of a detectable interaction in White and Non-Hispanics older adults may reflect greater heterogeneity in depression subtypes, better vascular health, or compensatory mechanisms that moderate WMH effects. These findings indicate that cerebrovascular pathology may contribute to cognitive decline that is influenced by both depression and cultural context. Future research should incorporate measures of depression severity and timing (e.g., early vs. late onset), alongside detailed vascular assessments and culturally informed psychosocial metrics, to better understand the mechanisms underlying these differences. In addition to larger sample sizes particularly for minority groups, longitudinal designs are needed to clarify how depression trajectories and WMH burden evolve over time across diverse populations.

## Data Availability

Data used in preparation of this article were also obtained from the National Alzheimers Coordinating Center (NACC, https://naccdata.org/) database

## Acknowledgments

The authors acknowledge use of Compute Canada (https://alliancecan.ca/en) resources for performing the image processing and WMH segmentations in the presented work. The NACC database is funded by NIA/NIH Grant U24 AG072122. NACC data are contributed by the NIA-funded ADRCs: P30 AG062429 (PI James Brewer, MD, PhD), P30 AG066468 (PI Oscar Lopez, MD), P30 AG062421 (PI Bradley Hyman, MD, PhD), P30 AG066509 (PI Thomas Grabowski, MD), P30 AG066514 (PI Mary Sano, PhD), P30 AG066530 (PI Helena Chui, MD), P30 AG066507 (PI Marilyn Albert, PhD), P30 AG066444 (PI David Holtzman, MD), P30 AG066518 (PI Lisa Silbert, MD, MCR), P30 AG066512 (PI Thomas Wisniewski, MD), P30 AG066462 (PI Scott Small, MD), P30 AG072979 (PI David Wolk, MD), P30 AG072972 (PI Charles DeCarli, MD), P30 AG072976 (PI Andrew Saykin, PsyD), P30 AG072975 (PI Julie A. Schneider, MD, MS), P30 AG072978 (PI Ann McKee, MD), P30 AG072977 (PI Robert Vassar, PhD), P30 AG066519 (PI Frank LaFerla, PhD), P30 AG062677 (PI Ronald Petersen, MD, PhD), P30 AG079280 (PI Jessica Langbaum, PhD), P30 AG062422 (PI Gil Rabinovici, MD), P30 AG066511 (PI Allan Levey, MD, PhD), P30 AG072946 (PI Linda Van Eldik, PhD), P30 AG062715 (PI Sanjay Asthana, MD, FRCP), P30 AG072973 (PI Russell Swerdlow, MD), P30 AG066506 (PI Glenn Smith, PhD, ABPP), P30 AG066508 (PI Stephen Strittmatter, MD, PhD), P30 AG066515 (PI Victor Henderson, MD, MS), P30 AG072947 (PI Suzanne Craft, PhD), P30 AG072931 (PI Henry Paulson, MD, PhD), P30 AG066546 (PI Sudha Seshadri, MD), P30 AG086401 (PI Erik Roberson, MD, PhD), P30 AG086404 (PI Gary Rosenberg, MD), P20 AG068082 (PI Angela Jefferson, PhD), P30 AG072958 (PI Heather Whitson, MD), P30 AG072959 (PI James Leverenz, MD).

## Consent Statement

Written informed consent was obtained from participants or their study partner

## Availability of data and materials

Data used in preparation of this article were also obtained from the National Alzheimer’s Coordinating Center (NACC, https://naccdata.org/) database, including the NACC Uniform Data Set (UDS), and MRI Data Set (Beekly et al., 2004; Besser, Kukull, Knopman, et al., 2018; Besser, Kukull, Teylan, et al., 2018). The image processing and WMH segmentation pipelines used are open source and available at https://github.com/VANDAlab/Preprocessing_Pipeline. The lobar atlas used to derive regional WMH metrics is also available at https://zenodo.org/records/7930159.

## Contributions

F.K, R.M, M.D, and C.M, were involved with the conceptualization and design of the work. F.K. and M.D. completed analysis and C.M, M.D, R.M, and F.K were involved with data interpretation.

F.K. organized figures. F.K. wrote the manuscript. and C.M, R.M, M.D, and F.K revised and approved the submitted version.

## Consent for publication

Not applicable.

